# An international study to investigate and optimise the safety of discontinuing valproate in young men and women with epilepsy: protocol

**DOI:** 10.1101/2024.06.16.24308993

**Authors:** Gashirai K. Mbizvo, Glen P. Martin, Matthew Sperrin, Laura J. Bonnett, Pieta Schofield, Iain Buchan, Gregory Y.H. Lip, Anthony G. Marson

## Abstract

Valproate is the most effective treatment for idiopathic generalised epilepsy. Currently, its use is restricted in women of childbearing potential owing to high teratogenicity. Recent evidence extended this risk to men’s offspring, prompting recommendations to restrict use in everybody aged <55 years.

This study will evaluate mortality and morbidity risks associated with valproate withdrawal by emulating a hypothetical randomised-controlled trial (called a “target trial”) using retrospective observational data. The data will be drawn from ∼250m mainly US patients in the TriNetX repository and ∼60m UK patients in Clinical Practice Research Datalink (CPRD). These will be scanned for individuals aged 16–54 years with epilepsy and on valproate who either continued, switched to lamotrigine or levetiracetam, or discontinued valproate between 2015–2018, creating four groups. Randomisation to these groups will be emulated by baseline confounder adjustment using g-methods. Mortality and morbidity outcomes will be assessed and compared between groups over five years, employing time-to-first-event and recurrent events analyses. A causal prediction model will be developed from these data to aid in predicting the safest alternative antiseizure medications.

Together, these findings will optimise informed decision-making about valproate withdrawal and alternative treatment selection, providing immediate and vital information for patients, clinicians and regulators.

## Background

Epilepsy impacts 70 million people worldwide.^1^ Valproate is one of the most effective drugs for treating epilepsy, outperforming any other in idiopathic generalised epilepsy, which affects a third of people with epilepsy (PWE).^2,3^ It was the most prescribed antiseizure medication (ASM) globally in the early 2000s.^4^ However, widespread evidence demonstrating foetal congenital malformations (11%) and neurodevelopmental disorders (30–40%) associated with prenatal valproate exposure led to guidance changes between 2015–2018.^5^ These prohibited valproate prescribing in women of childbearing potential unless other ASMs had failed, subsequently halving prescriptions in women.^4,5^

Valproate remains the first-line ASM for men with newly diagnosed generalised epilepsy in most countries.^2,3^ In the UK, regulators recently considered valproate’s known risks of impaired male fertility, emerging pre-clinical evidence of transgenerational risks with prenatal exposure, and animal studies suggesting adverse effects on testes.^5,6^ In response, they announced a phased implementation (from January 2024) of recommendations restricting valproate prescribing for both men and women aged <55 years unless other ASMs have failed.^5,6^ Similar restrictions are being considered in other countries.^7-9^ Together, they are likely to result in withdrawal of valproate from a substantial number of men already taking it, as has been the case for women.^4,5^

To support informed and safe decision-making,^10^ it is vital to provide these young men and women and their clinicians with comprehensive information on the safety of valproate withdrawal. Whether there are any potential risks of personal harm or morbidity (including hospital admissions, injuries, or death) associated with valproate withdrawal has not yet been quantified for either men or women on valproate.^10-17^ Existing studies have focused on the group of PWE in remission from seizures for ≥2 years, usually showing seizure relapse in generalised epilepsy. They have tended to be single-centre or small, looking at seizures alone or assessing women alone.^11-17^ A valproate decision support tool is available from NHS England for patients and clinicians (www.t.ly/NFi5m), designed by our centre.^10^ Figures for the risks of teratogenicity on valproate are provided on page 4 of the tool as 8–37 per 100 babies.^10^ However, on the same page, the “risks to you” section from withdrawing from valproate and switching to another ASM remains devoid of any figures,^10^ illustrating the insufficiency of current evidence to address this question.

The rise of digital care records has transformed treatment evidence generation by enabling both immediate and extensive analysis of national and international cohorts.^18^ This approach overcomes limitations in sample size, cost, and time associated with completing prospective studies while also facilitating exploration of previously unexamined patient outcomes. TriNetX,^19^ the world’s largest real-world electronic health data platform, harbours 70 billion date- and patient-indexed electronic clinical observations from ∼250m patients across 19 countries, predominantly in America but also Europe, Middle East, Africa, and Asia.^18,19^

We recently published a study of 8,991 men and 5,243 women aged 16–54 with epilepsy taking valproate in TriNetX. This was a survival analysis assessing 5-year mortality and morbidity in those withdrawn from valproate between 01/06/2015–01/06/2018 (regardless of whether switched to another ASM), with propensity-matched comparisons to those who remained on valproate. Results suggested substantially increased risks of emergency department (ED) attendances, hospital admissions, falls, injuries, burns and new-onset depression in both men and women withdrawn from valproate.^20^ However, these were aggregate data designed to scope potential trends and generate hypotheses, not to probe causal links.

To progress the field, we propose a comprehensive study examining mortality and morbidity outcomes in individuals who either continued valproate, switched to lamotrigine or levetiracetam, or discontinued valproate. Employing a novel approach known as target trial emulation,^21,22^ this study will replicate a hypothetical randomised-controlled trial (RCT) using person-level retrospective observational data from both TriNetX and Clinical Practice Research Datalink (CPRD, which encompasses 60m UK patients).^23^ Target trial emulation minimises confounder and selection biases in observational studies by statistically emulating randomisation at baseline (time 0).^21^ Individuals are then analysed based on the treatment strategy aligned with their data. Results are available quickly and shown to mirror those of an RCT.^21^ This offers a cost-effective and immediate alternative to conducting an RCT;^21^ crucial given the urgency for information on valproate withdrawal safety amidst ongoing regulatory changes.^24,25^ Additionally, the emulation facilitates creation of causal prediction models.^26^ These are developed using observed data alongside causal theory and assumptions to calculate predicted risk under different hypothetical treatment conditions, allowing “what if” questions to be addressed for each potential (counterfactual) outcome.^26^ This facilitates actionable prevention.^26^

### Aims

1. Identify whether there are any differences in mortality or morbidity outcomes between remaining on valproate compared with valproate withdrawal and switch to lamotrigine, levetiracetam, or no new ASM.
2. Develop and validate a causal prediction model that clinicians and patients could use when planning to withdraw from valproate to predict which of these alternative ASMs would have the lowest mortality and morbidity, taking baseline clinical and demographic characteristics into account.

### Hypotheses

1. Some morbidity or mortality outcomes will be increased when valproate is withdrawn, regardless of whether switched to lamotrigine, levetiracetam, or no new ASM;
2. The causal prediction model developed will be able to predict which alternative ASMs are safest.

### Benefits to PWE

This study will benefit PWE as substantial numbers of young men and women may soon be invited by their clinicians to come off valproate owing to both current and new regulatory changes.^5,6^ Whilst this is justified in terms of preventing teratogenic harm, the patients will wish to know what the risks of withdrawing from valproate are for their own health.^24,25^ This study will provide immediate evidence to help doctors answer that question.^10^ It will also help patients make safer decisions over switching to alternative ASMs by generating the first ever causal prediction tool for this purpose.^26^ This will be the largest study of valproate withdrawal to date, the first to assess risks in men and first to assess multiple outcomes beyond seizures alone for both men and women.^11-17^ These outcomes will be collaboratively chosen with PWE through public engagement strategies, ensuring their relevance and impact on those most affected. Finally, mortality prediction is highlighted as a key recommendation for epilepsy research by NICE,^27^ meaning the causal prediction model generated by this study also addresses an established research need for PWE.

## Methods

### Public involvement

This study protocol (including outcome selection) was co-developed with a members of the public with lived experience of epilepsy, caring for a person with epilepsy, or taking valproate.

**Addressing Aim 1:** target trial emulation

### Target trial design

A hypothetical target trial will be emulated in which young men and women taking valproate would be randomised to either continue valproate, switch to lamotrigine or levetiracetam, or discontinue valproate.^21^ The following mortality and morbidity outcomes would be assessed over the subsequent five years:

– Deaths (all-cause/epilepsy-related);
– ED/hospital admissions (all-cause/epilepsy-related);
– Seizures (symptom codes);
– Falls;
– Injuries;
– Burns;
– Aspiration pneumonia;
– Manic/bipolar episodes;
– New-onset depression;
– Self-harm/suicide;
– A composite morbidity index incorporating seizures, injuries, admissions, depression.

Intention-to-treat (ITT) and per-protocol effects would be assessed between groups.^21^ Analysis would take a time-to-first-event approach (Cox-proportional hazards model) for deaths, new-onset depression and composite morbidity. A recurrent events approach (e.g., Prentice-Williams-Peterson Total Time Model) would be taken for the remaining outcomes.

### Target trial emulation using observational data from TriNetX and CPRD

#### Eligibility criteria

Anonymised, person-level, electronic health data will be extracted from TriNetX and CPRD, linked across primary care, ED, inpatients, outpatients, and mortality subsets. Included will be men and women aged 16–54 years with ≥1 epilepsy disease or symptom code between 01/06/2015–01/06/2018 and ≥1 valproate prescription instance within the preceding 6 months and 6–12 months before the first epilepsy disease or symptom code to appear between 01/06/2015–01/06/2018. TriNetX and CPRD use ICD-10-CM and SNOMED codes, respectively. The coding strategy combining disease and symptom codes with valproate, as used our published study successfully,^20^ was shown to accurately identify epilepsy cases in our prior work.^28,29^

### Treatment armsxs

1. Withdrawn valproate and commenced lamotrigine
2. Withdrawn valproate and commenced levetiracetam
3. Withdrawn valproate and no new ASM commenced
4. Continues valproate

### Treatment assignment

Baseline (time 0) will be defined as when valproate dose reduction commences (treatment arms 1–3) or prescription of valproate at a matched time (arm 4). Eligible individuals are assigned at baseline to the treatment strategy that their data are consistent with.^21^ To emulate randomisation, adjustment is made for baseline confounders using Robins’ generalised methods (g-methods, e.g., inverse probability weighting),^21^ considering age, sex, ethnicity, deprivation, type of epilepsy (using G40 codes), baseline seizure frequency (symptom codes recorded over preceding 12 months), alcohol excess, developmental delay, Cambridge Multimorbidity Score, length of valproate treatment, valproate dose, and ASMs used other than valproate. The final set of covariates required to adjust for confounding will be chosen using a causal directed acyclic graph.^21^

### Outcomes

These are listed in the target trial design section and derived from our prior work,^1,20,30,31^ and public consultation. All coded events will be recorded for flexible categorical or continuous analyses, as needed.

#### Causal estimand

Assesses impact of treatment arms 1–3 vs. 4 on outcome (per-protocol assumption as observational data limits ITT assessment).^21^

### Start and end of follow-up

Beginning at baseline (time 0), as defined above, and followed-up till outcome, censoring (including deviation from assigned treatment), or five years.

#### Statistical analysis

mirrors target trial section.

**Addressing Aim 2:** causal prediction modelling

A causal prediction model will be created using Cox-proportional hazards of time-to-first-event in a causal framework.^26^ The predicted outcome will be a composite personal harms risk combining epilepsy-related deaths, seizures, injuries, admissions, and depression. Predictors for definite inclusion in the final model will be treatment arms 1–4. Candidate predictors considered in addition will be taken from the list of baseline confounders adjusted in the target trial emulation, alongside any additional variables considered to influence outcome but not treatment assignment. *A priori* clinical expertise and data-driven variable selection will refine the predictor variable list for modelling.^30^ Penalisation methods (e.g., *lasso*), will be used to combine variable selection with shrinkage (reducing overfitting risks).

The model will be developed in TriNetX to help ensure it is internationally generalisable.^32^ External validation will be undertaken in CPRD, facilitating subsequent clinical implementation.^32^ Internal validation within TriNetX will involve 1,000 bootstrap analysis samples. Nagelkerke’s R^2^ and Brier Scores will estimate overall model performance. Calibration will be estimated with calibration-in-the-large, calibration slope, and flexible calibration plots, and discrimination with C-index. Calibration and discrimination will be reassessed within CRPD to externally validate model performance.^32^ Counterfactual prediction will be embedded into the model development,^26^ allowing patients and clinicians to answer clinically meaningful questions such as “at the point of proposed valproate withdrawal, accounting for baseline characteristics, what is the risk of epilepsy-related death or composite morbidity for levetiracetam? And what is it for lamotrigine?”

## Sample size calculation

See table 1.

**Table 1:** Sample size calculation.

| Outcome: Emergency department attendance (a global measure of morbidity and a good predictor of mortality)<br>Data source: Mbizvo et al.<sup>20</sup> |  |  |  |
| --- | --- | --- | --- |
| Target trial emulation phase |  | Prediction model phase |  |
| **Method** | `# Proportions from the pilot study p1 <- 2977 / 8021 # valproate withdrawn group p2 <- 2471 / 8021 # group remaining on valproate # Set significance level and power alpha <- 0.05 power <- 0.80 # Z-values for two-tailed test z_alpha <- qnorm(1 - alpha / 2) z_power <- qnorm(power) # Calculate sample size n <- ceiling((z_alpha + z_power)^2 * ((p1 * (1 - p1)) + (p2 * (1 - p2))) / (p1 - p2)^2) # Display the result cat("Sample size needed for the target trial emulation study:", n, "\n")` | **Method** | Sample size criteria from our previous work for developing a time-to-event prediction model to estimate the minimum required sample size for our model development are used.<sup>33</sup> |
| **Result** | The minimum sample size in the target trial emulation study is 881. | **Assumptions** | (i) We target an overall shrinkage factor of 0.9.<br>(ii) We assume the outcome rate to be 0.068, calculated using the 5,448 events that occurred out of 16,042 patients within the pilot study, with a median survival time of 5 years.<br>(iii) The performance of the model would achieve 15% of the maximum possible cox-snell R-squared for the given outcome rate.<br>(iv) We would consider 20 candidate predictor parameters within the model development process.<br>**Outcome based these assumptions**<br>The minimum sample size for model development is 1,204 observations. |
| **Feasibility** | The TriNetX sample, derived from the pilot study, captures approximately 21,000 people with epilepsy prescribed valproate between 2015–2018, with half subsequently withdrawn. This is sufficient. I have also undertaken a CPRD feasibility search, identifying 29,332 men and 15,315 women with epilepsy prescribed valproate in between 2010–2021, which is also sufficient. |  |  |

## Contingency planning

Studying both TriNetX and CPRD allows external cross-validation of findings between the two databases, enhancing generalisability whilst providing a contingency study cohort should one dataset encounter limitations.

### Pathways to impact

The study will be published open-access in peer-reviewed journals and shared with regulators, charities, and PWE. Following our guide,^34^ we will explore various presentation formats for the externally validated prediction model including points-based systems, graphical score charts, and nomograms, as informed by PWE at public engagement workshops and clinicians at scientific conferences. An interactive version of the final causal prediction model will be deposited online in a public repository (as was did for the mortality prediction tool we developed previously: https://seds-tool.github.io/seds), making it globally accessible. We did similar with the CHA<sub>2</sub>DS<sub>2</sub>-VASc score we created and deposited on MDCalc.com (www.t.ly/poRmb): now the most widely incorporated prediction tool for stroke prevention in atrial fibrillation globally, embedded in NICE guidelines (www.t.ly/d0d3W). We will also add the model to the Civic Data Cooperative (https://civicdatacooperative.com/about) Digital Commons (https://github.com/civicdatacoop) to facilitate integration into health system core infrastructure.

Colleagues at our centre who created NHS England’s valproate decision support tool (www.t.ly/NFi5m) are in support of figures generated in addressing Aim 1 being added to the medication change “risks to you” section on page 4, alongside including a link to the online version of the causal prediction model generated in addressing Aim 2.^10^

## Ethics and dissemination

The University of Liverpool Research Ethics Committee has provided formal ethics review exemption for this study as it does not involve human participants, human tissue or personal data (REC Ref. 14455). Dissemination is outlined in the pathways to impact section.

## Data management plan

Data will be curated through University of Liverpool’s (UoL) Active Data Storage -a centralised, secure, supported data storage facility with multiple layers of protection. Data are replicated between two secure physical locations and backed up regularly. A regular tape backup is made to a third physical location, and segregated from the public network both physically and logically. Data are encrypted in transit using SSL.

We will use a public repository (www.github.com) to make all diagnostic and outcome coding algorithms, metadata, and R analysis scripts used publicly available, facilitating external replication and adaptation.

All data storage and use will comply with legal obligations (including GDPR) and UoL’s Research Data Management Policy.

## Study status and timeline

Funding has been received from the Epilepsy Research Institute and the Academy of Medical Sciences to proceed with this study. Permissions to access study data are being sought and PhD students to help complete the study are being recruited. We plan to complete the study within 36 months of protocol submission/publication..

## Data Availability

No datasets were generated or analysed during the current study. All relevant data from this study will be made available upon study completion.

## Acknowledgements

We thank our Public Advisors (including Sean Barnes and Steph Tomlinson) for support with this study, and NIHR Applied Research Collaboration North West Coast for providing access to Public Advisors.

